# Is annual vaccination best? A modelling study of influenza vaccination strategies in children

**DOI:** 10.1101/2021.09.22.21263935

**Authors:** Kylie E. C. Ainslie, Steven Riley

**Affiliations:** School of Public Health, Imperial College London, London, UK; MRC Centre for Global Infectious Disease Analysis, Department of Infectious Disease Epidemiology, School of Public Health, Imperial College London, London, UK

## Abstract

**Introduction:** Annual vaccination of children against influenza is a key component of vaccination programs in many countries. However, past infection and vaccination may affect an individual’s susceptibility to infection. Little research has evaluated whether annual vaccination is the best strategy. Using the United Kingdom as our motivating example, we assess the impact of different childhood vaccination strategies, specifically annual and biennial (every other year), on attack rate and expected number of infections.

**Methods and Findings:** We present a multi-annual, individual-based, stochastic, force of infection model that accounts for individual exposure histories and disease/vaccine dynamics influencing susceptibility. We simulate birth cohorts that experience yearly influenza epidemics and follow them until age 18 to determine attack rates and the number of childhood infections. We perform simulations under baseline conditions, with an assumed vaccination coverage of 44%, to compare annual vaccination to no and biennial vaccination. We relax our baseline assumptions to explore how our model assumptions impact vaccination program performance.

At baseline, we observed more than a 50% reduction in the number of infections between the ages 2 and 10 under annual vaccination in children who had been vaccinated at least half the time compared to no vaccination. When averaged over all ages 0-18, the number of infections under annual vaccination was 2.07 (2.06, 2.08) compared to 2.63 (2.62, 2.64) under no vaccination, and 2.38 (2.37, 2.40) under biennial vaccination. When we introduced a penalty for repeated exposures, we observed a decrease in the difference in infections between the vaccination strategies. Specifically, the difference in childhood infections under biennial compared to annual vaccination decreased from 0.31 to 0.04 as exposure penalty increased.

**Conclusion:** Our results indicate that while annual vaccination averts more childhood infections than biennial vaccination, this difference is small. Our work confirms the value of annual vaccination in children, even with modest vaccination coverage, but also shows that similar benefits of vaccination can be obtained by implementing a biennial vaccination program.

**Author summary:** Many countries include annual vaccination of children against influenza in their vaccination programs. In the United Kingdom, annual vaccination of children aged of 2 to 10 against influenza is recommended. However, little research has evaluated whether annual vaccination is the best strategy, while accounting for how past infection and vaccination may affect an individual’s susceptibility to infection in the current influenza season. Prior work has suggested that there may be a negative effect of repeated vaccination. In this work we developed a stochastic, individual-based model to assess the impact of repeated vaccination strategies on childhood infections. Specifically, we first compare annual vaccination to no vaccination and then annual vaccination to biennial (every other year) vaccination. We use the UK as our motivating example. We found that an annual vaccination strategy resulted in the fewest childhood infections, followed by biennial vaccination. The difference in number of childhood infections between the different vaccination strategies decreased when we introduced a penalty for repeated exposures. Our work confirms the value of annual vaccination in children, but also shows that similar benefits of vaccination can be obtained by implementing a biennial vaccination program, particularly when there is a negative effect of repeated vaccinations.

## Introduction

Influenza is a significant cause of morbidity and mortality annually [1], which is likely to cause substantial waves of infection as COVID-19 social distancing is relaxed. Pre-COVID-19, 20% to 30% of children were infected with influenza compared to only 5% to 10% of adults, annually [2]. The World Health Organization lists children under the age of 5 years old as a target group for influenza vaccination [3] because more than 100,000 children under five die of influenza each year, and attack rates in children are fourto five-fold higher in children compared to adults [2]. Additionally, influenza infection in children can lead to complications and severe disease with the highest incidence of influenza-related hospitalisations in young children [4]. Due to the disease burden of influenza, the United Kingdom began offering the influenza vaccine to all primary school aged children (those aged 6-10 years) in England in the winter of 2019/2020 for the first time [5]. This group was also targeted because school-aged children are thought to drive influenza transmission [6]. Prior modelling work has shown that adopting a transmission-based approach to vaccination can reduce the spread of influenza by targeting groups that drive transmission (such as children) [7].

Modelling is an important tool that can aid policy decisions surrounding vaccination programs by exploring the impacts of different strategies and scenarios [8]. Unlike other childhood vaccines, the influenza vaccine must be re-administered each year due to continuous changes in the virus [9, 10]; therefore decisions to implement a recurring influenza vaccination policy must be weighed against other considerations, such as economic cost. Many prior modelling studies have focused on quantifying the impacts of childhood vaccination programs [8, 11–16], including studies that have specifically assessed the impact of childhood vaccination programs in England [7, 12, 13]. Overwhelmingly, these studies have found both direct and indirect benefits of childhood vaccination programs. Additionally, studies have found that prevention of influenza infection and subsequent disease by vaccination is a cost-effective method of preventing influenza [17, 18]. While the specific assumptions and models vary across these studies, they all focus on annual vaccination programs and do not account for the potential negative effects of repeated exposures.

It is still unclear whether repeated influenza vaccination results in negative effects whereby individuals who receive multiple vaccines over a short period of time have increased risk of infection compared to individuals who received a vaccine only in the current season. The first report of the potential negative effects of repeated annual vaccination was published in a series of papers describing influenza outbreaks at a British boarding school from 1972 to 1976 [19–21]. Repeated influenza vaccination has received growing interest [22] and observational and modelling studies have shown that repeated annual vaccination may result in less protection against influenza infection [21, 23–28]. This phenomenon may be due to an individual’s immune system generating the strongest response to their first exposure and then less strong responses to subsequent exposures of a similar type [29]. Alternatively, the increased risk of infection after repeated vaccination could be due to negative interference from closely related vaccine strains such that the response to the second vaccine strain is partially eliminated by pre-existing cross-reactive antibodies produced in response to the first vaccine strain [23]. The precise mechanism whereby reduced protection after repeated vaccination remains unclear.

Here, we assessed how different frequencies of vaccination may impact childhood infections while also accounting for potential negative impacts of repeated exposures. Using a multi-annual, individual-based, stochastic, force of infection model we evaluate the impact of different childhood vaccination strategies. A strength of our approach is that our individual-based model accounts for each individual’s exposure history and disease/vaccine dynamics influencing susceptibility, namely antigenic drift, vaccine match, and waning. This work could have impact on future policy surrounding repeated vaccination programs, but the focus of this paper is not on changing policy, but rather on quantifying potential benefits of 1) current annual vaccination strategies in children and 2) alternative strategies for those countries where annual vaccination programs are not feasible. We use the vaccination program in the UK as our motivating example. We first assess annual vaccination programs in young and school-aged children (aged 2-10 years) compared to no vaccination program. We next assess the benefits of a biennial (every other year) vaccination program compared to an annual program. We quantify the performance of each vaccination program over many seasons as the expected number of infections children experience during childhood.

## Materials and methods

### Model

Influenza A is a rapidly evolving virus [30] allowing for numerous infections over time [31]. Many countries recommend frequent vaccination [5, 32, 33], thus the number of exposures to influenza in a person’s lifetime may be many. In order to model the effects of repeated exposures over an individual’s lifetime, we have developed a multi-annual, individual-based, stochastic model of infection and vaccination. Our model incorporates three main components: 1) viral evolution, specifically antigenic drift of the infecting virus over time, 2) vaccine kinetics influencing the amount of protection conferred by the vaccine, namely vaccine update, waning, and take, and 3) individual level characteristics, such as age and prior exposure history. All three components are then used to inform an individual’s susceptibility to infection at each time point (here, considered to be one year). Our model is implemented in a series of steps at each time point:

#### 1. Determine the amount of antigenic distance

Within each year, we first determine the amount of antigenic drift from the previous year’s virus. We incorporate viral drift into the model as a random process. Each year the antigenic distance from the previous year’s circulating strain is drawn from an exponential distribution with rate 1 [34, 35].

#### 2. Vaccine update

Influenza vaccines are updated frequently to account for antigenic drift [36]. However, they are only updated when there has been significant evolution of the viral strains [37]. The cumulative antigenic distance since the last vaccine update determines when the next vaccine update will occur. When the cumulative sum of the antigenic distance since the last vaccine update reaches a threshold (for example, 3 antigenic units) the vaccine is updated (Fig 1A).

**Fig 1.**
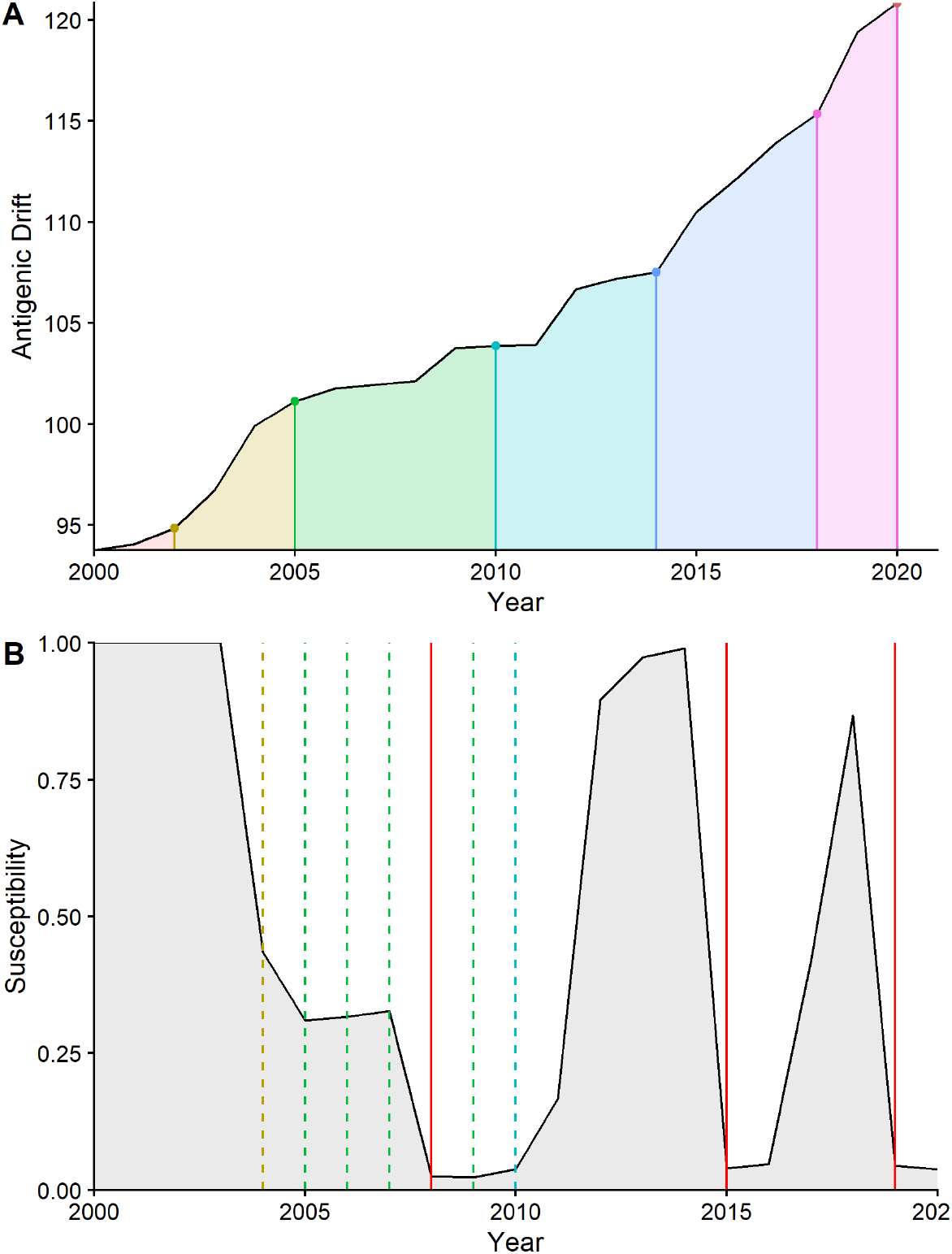
A. Plot of antigenic distance (black line) with corresponding vaccine updates (coloured dots). Shaded regions below the black line indicate the different vaccines used for the period of time between updates. B. an individual susceptibility profile indicating their susceptibility over time (black line) and exposures: infections (red line) and vaccinations (dashed lines). The colour of the dashed lines indicates the vaccine that was used and corresponds to the colours in A. This is an example profile that assumes annual vaccination.

a. **Determining amount of protection conferred by the vaccine**. After determining whether the vaccine strain is updated, the amount of protection conferred by the vaccine in each year is determined. When the vaccine matches the circulating strain, it provides provides 70% protection (i.e., vaccine efficacy is 70%). In years between updates (i.e., years in which the vaccine and circulating strain do not match), vaccine efficacy is reduced proportional to the antigenic distance between the vaccine and circulating strains.

#### 3. Vaccinate

Each individual may be vaccinated. Depending on the vaccine strategy, individuals may be vaccinated every year or every other year. Probability of vaccination is correlated such that an individual who was vaccinated in the previous cycle (i.e., the previous year under an annual vaccination strategy or two years previously under biennial vaccination strategy) has a higher probability of being vaccinated in the current cycle compared to someone who was not vaccinated in the previous cycle. When accounting for prior vaccination, we only consider the most recent previous cycle, not an individual’s entire vaccination history. Each individual’s vaccination status is recorded each year and used to inform their susceptibility in subsequent years.

a. We assume a leaky vaccine model [38]. In other words, we assume that everyone who receives a vaccine is partially protected from infection.
b. Immunity from vaccination is assumed to wane a fixed percentage each year, although there is some evidence that live-attenuated influenza vaccines may provide protection for longer [39]. Waning is modelled as a separate term, so that it is distinct from antigenic drift of the circulating strain away from the vaccine strain.

#### 4. Susceptibility

An individual’s susceptibility at any time depends on their exposure history (both from infection and vaccination). Each individual is born naive (with a susceptibility of 1) and their susceptibility decreases as the individual accumulates exposures (Fig 1B). We use the haemagglutination-inhibiting antibody protection curve described by Coudeville *et al*. and the parameter estimates from their ALL model (*α* = log(2.844), *β* = 1.299) [40] to determine an individual’s susceptibility to subsequent infection based on the antigenic distance between circulating and exposing viruses. When an individual is infected their susceptibility is assumed to be 0 immediately following infection and then increases as the exposing virus drifts. We accomplish this by setting their log titre to the value that represents full protection under the Coudeville model (log(200) = 5.30). As the virus drifts, the value of log titre decreases with a one to one relationship to the antigenic distance of the drifting virus. The new log titre value is then used to recalculate the individual’s susceptibility along the protection curve.

In the event an individual is vaccinated and they have no prior immunity from infection, their susceptibility becomes 1 minus the protective effect of the vaccine and then increases as immunity from the vaccine wanes and/or the virus drifts away from the vaccine strain. When an individual has immunity both from prior infection and vaccination their susceptibility at time *t* is their immunity from infection multiplied by their protection from vaccination. For example, consider an individual who was infected in year 2 and vaccinated in year 3. Their susceptibility in year 2 drops to 0 following infection (i.e., they are 100% immune to the infecting virus). In year 3, the virus drifts by 5% and the individual is vaccinated with a vaccine that is a perfect match to the circulating strain, thus conferring 70% protection. Therefore, the individual’s susceptibility is the product of the protection from both previous infection and vaccination, thus their susceptibility is 1.5% ((1 − 0.95) *×* (1 − 0.7) = 0.015).

#### 5. Infection

An individual may become infected with probability equal to the annual force of infection multiplied by an individual’s susceptibility at the current time point. Each individual’s infection status is recorded for every year and used to inform their susceptibility in subsequent years.

#### 6. Exposure penalty

Current scientific theory suggests that an individual’s immune system responds strongest to their first exposure and then responds less strongly to subsequent exposures of a similar type [29], such as multiple infections with influenza. This theory has been termed original antigenic sin [29] and may explain why recent observational studies of influenza vaccine effectiveness have found a detrimental affect of repeated influenza vaccination [26–28, 41]. To include this possible immunological mechanism in our model, we include an exposure penalty to account for decreased immune response to subsequent infections. After an individual is exposed, either by infection or vaccination, their maximum immunity (originally, 100%) decreases by ϵ, where ϵ = (0, 0.1). In other words, an individual’s minimum susceptibility increases with every exposure. An example can be seen in Fig 1B, where the first time the individual becomes infected (left-most red line) their susceptibility drops to 0. The second and third times they become infected (center and right-most red lines), their susceptibility is larger than zero due to exposures (vaccinations are represented as dashed lines) that occurred before the subsequent infections.

### Simulation

To determine how different vaccination strategies impact the number of infections children get during childhood (here, defined as between birth and 18 years old), we simulated annual influenza outbreaks from 1918 to 2028. We simulated a naive population of 30,000 individuals aged 0 to 79. We assume individuals die at age 80 and leave the population, and the birth rate is equal to the death rate, that is, every deceased person is replaced by a naive person. Pandemic influenza is introduced into the population in the first year (with a force of infection of 0.4), followed by annual seasonal epidemics with a constant force of infection (0.2) for the remaining years. We assume an individual can only be infected once a year and that only one strain is circulating. We perform a sensitivity analysis in which the force of infection varies from year to year. The time-varying force of infection is drawn from a logit normal distribution with distribution parameters *µ* = −1.49 and *σ* = 0.6, corresponding to a mean of 0.20, a standard deviation of 0.09 and a coefficient of variation of 46% [42].

Vaccination is introduced into the population in year 2000. Vaccination coverage is determined by age. In children, we test several different vaccination strategies. Current UK vaccination policy is to annually vaccinate children aged 2 to 10 years [5]. Thus, we implemented the following vaccination strategies in children: 1) no vaccination; 2) annual vaccination from ages 2 to 10; 3) biennial (every other year) vaccination from ages 2 to 10. We also perform a sensitivity analysis in which vaccination occurs from ages 2 to 16.

Within each simulation we identified birth cohorts beginning in 2000 until 2010, and followed each cohort until the cohort members reached age 18. We varied our model parameters (vaccination coverage, vaccine take, the amount of vaccine waning, exposure penalty, correlation of vaccination, and vaccine effectiveness) across simulations and performed 1000 simulations for each set of parameter values. We began with a baseline scenario with parameter values most favourable to the current annual vaccination strategy and determined the average number of childhood infections from each vaccination strategy. For this baseline scenario we assumed a vaccine efficacy of 70%, vaccination coverage of 44% (based on current UK estimates in children [43]), complete waning of vaccine-related protection after one year, 100% vaccine take, no exposure penalty, and a correlation of repeat vaccination of 90%. We then varied our parameter values using Latin hypercube sampling [44] to determine how the values of our model parameters influenced the difference in childhood infections across vaccination strategies. The parameters were varied within the following ranges: vaccine effectiveness = (0, 1), vaccination coverage = (0, 0.5), waning = (0, 1), take = (0.5, 1), exposure penalty = (0, 0.1), correlation of vaccination = (0, 1). Average childhood infections were determined across cohorts for each simulation and bootstrapping was used to obtain 95% confidence intervals of average number of childhood infections across simulations. All simulations were performed in R 4.0.2 [45].

The open source package `morevac` is available from GitHub (https://github.com/kylieainslie/morevac). The reader can run the model with different assumptions (parameter values) if they choose.

## Results

We used the model to assess the potential benefits of long-term annual repeated childhood vaccination in terms of annual attack rates and expected number of infections up to the age of 18 (Fig 2, Table 1). At ages with the lowest rates of infection, annual attack rates under the annual vaccination strategy were 2-fold lower compared to the no-vaccination strategy (Fig 2A). However, immediately after the oldest age of vaccination (11 years), we observed an increase in attack rate to a level greater than that expected from the no vaccination strategy. We observed the same pattern in expected number of childhood infections (Fig 2B). Overall, no vaccination resulted in 2.63 (95% confidence interval: 2.62, 2.64) infections up to the age of 18 while under annual vaccination the expected number of childhood infections decreased to 2.07 (2.06, 2.08) (Table 1). However, when we look at the ages in which children get infected we see that during the vaccination program (ages 2-10) there is a more pronounced difference in numbers of infections between vaccination strategies. We observed more than a 50% decrease in infections under an annual vaccination program for those who were vaccinated at least half the time (i.e., received at least 5 vaccinations between the ages of 2-10) compared to no vaccination strategy (Fig 2, Table 1).

**Fig 2.**
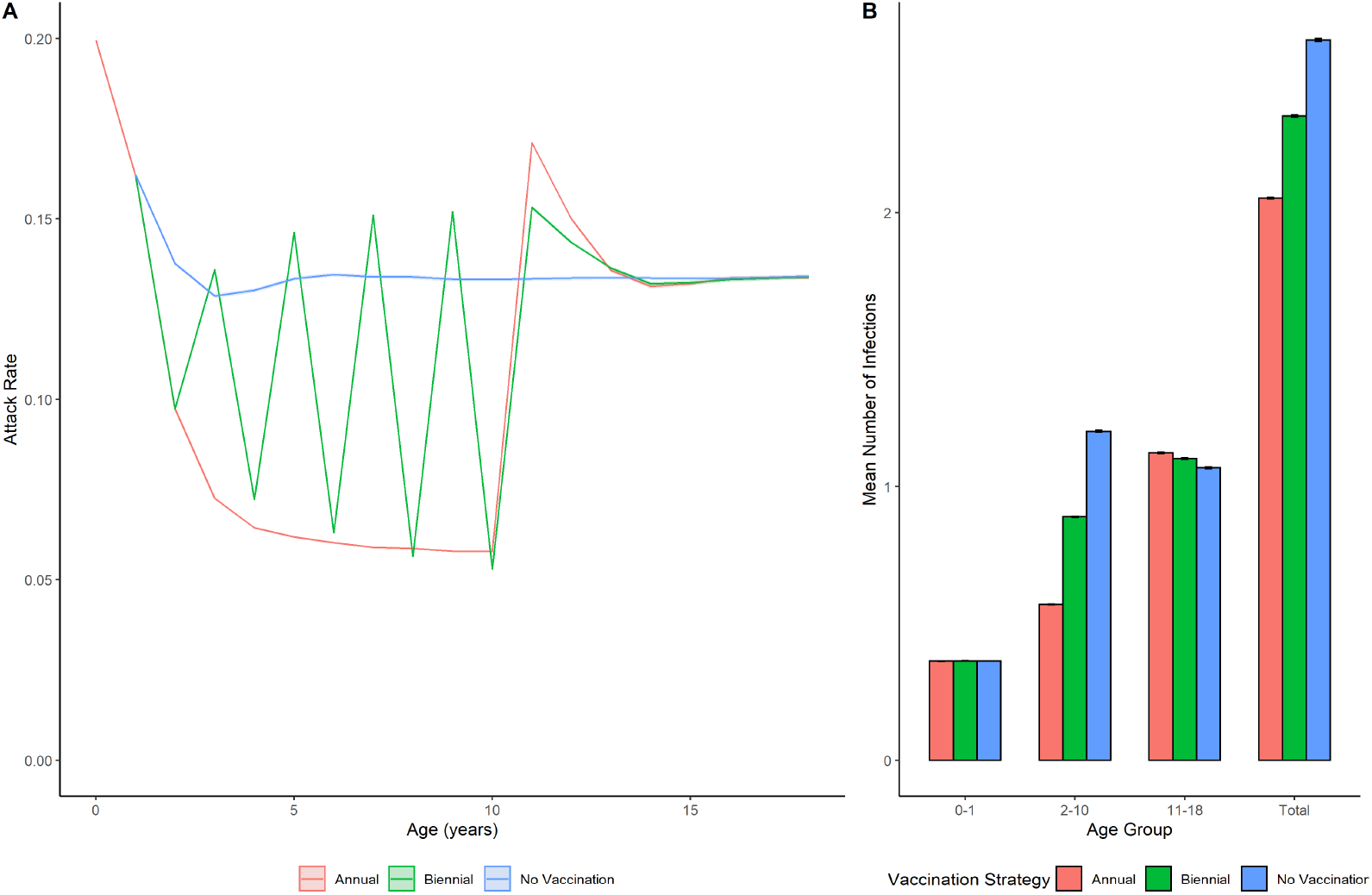
Average annual attack rates (A) and expected number of infections (B) in birth cohorts under different vaccination strategies: annual (red), biennial (green), and no vaccination (blue) under baseline assumptions. Lines/shading indicate mean 95% percentiles from 1000 simulations. Children aged 2 to 10 years were vaccinated.

**Table 1.** Number of expected childhood infections by age group under baseline assumptions comparing no vaccination strategy to annual and biennial vaccination programs in those who have received a vaccine at least half the time (≥ 5 vaccinations under annual, ≥ 3 vaccinations under biennial). Vaccination occurs during ages 2-10.

| Vaccination Strategy | Age Group | Infections (95% CI) |
| --- | --- | --- |
| Annual | Total | 2.05 (2.05, 2.06) |
|  | 0-1 | 0.362 (0.361, 0.362) |
|  | 2-10 | 0.568 (0.567, 0.570) |
|  | 11-18 | 1.12 (1.12, 1.13) |
| Biennial | Total | 2.35 (2.35, 2.36) |
|  | 0-1 | 0.362 (0.362, 0.363) |
|  | 2-10 | 0.889 (0.887, 0.892) |
|  | 11-18 | 1.10 (1.10, 1.10) |
| No Vaccination | Total | 2.63 (2.63, 2.64) |
|  | 0-1 | 0.362 (0.362, 0.363) |
|  | 2-10 | 1.20 (1.20, 1.20) |
|  | 11-18 | 1.07 (1.06, 1.07) |

The benefit from repeated annual vaccination was sensitive to our assumption about the immunological exposure penalty. To better understand the effect of exposure penalty, we plotted the attack rates (Fig 3) and determined the number of childhood infections (Table 1) for different values of exposure penalty under the baseline scenario. For plausible values of the exposure penalty, we observed substantial reductions in the benefits of repeated annual vaccination. We found that the benefit of annual vaccination decreased as exposure penalty increased, specifically, we saw an increase in attack rate in later years of childhood as the number of vaccinations individuals received increased (Fig 3). The expected number of childhood infections under annual vaccination increased with increasing exposure penalty from 2.07 (2.06, 2.08) infections with no exposure penalty to 2.48 (2.47, 2.49) infections with an exposure penalty of 0.1 (Table 2). However, even at our maximum value of exposure penalty, we still observed a reduction in childhood infections under annual vaccination compared to no vaccination (2.48 (2.47, 2.49) and 2.64 (2.63, 2.65), respectively). Our results were robust to the value of force of infection when we allowed force of infection to vary by year (Fig. S3, Table S1).

**Fig 3.**
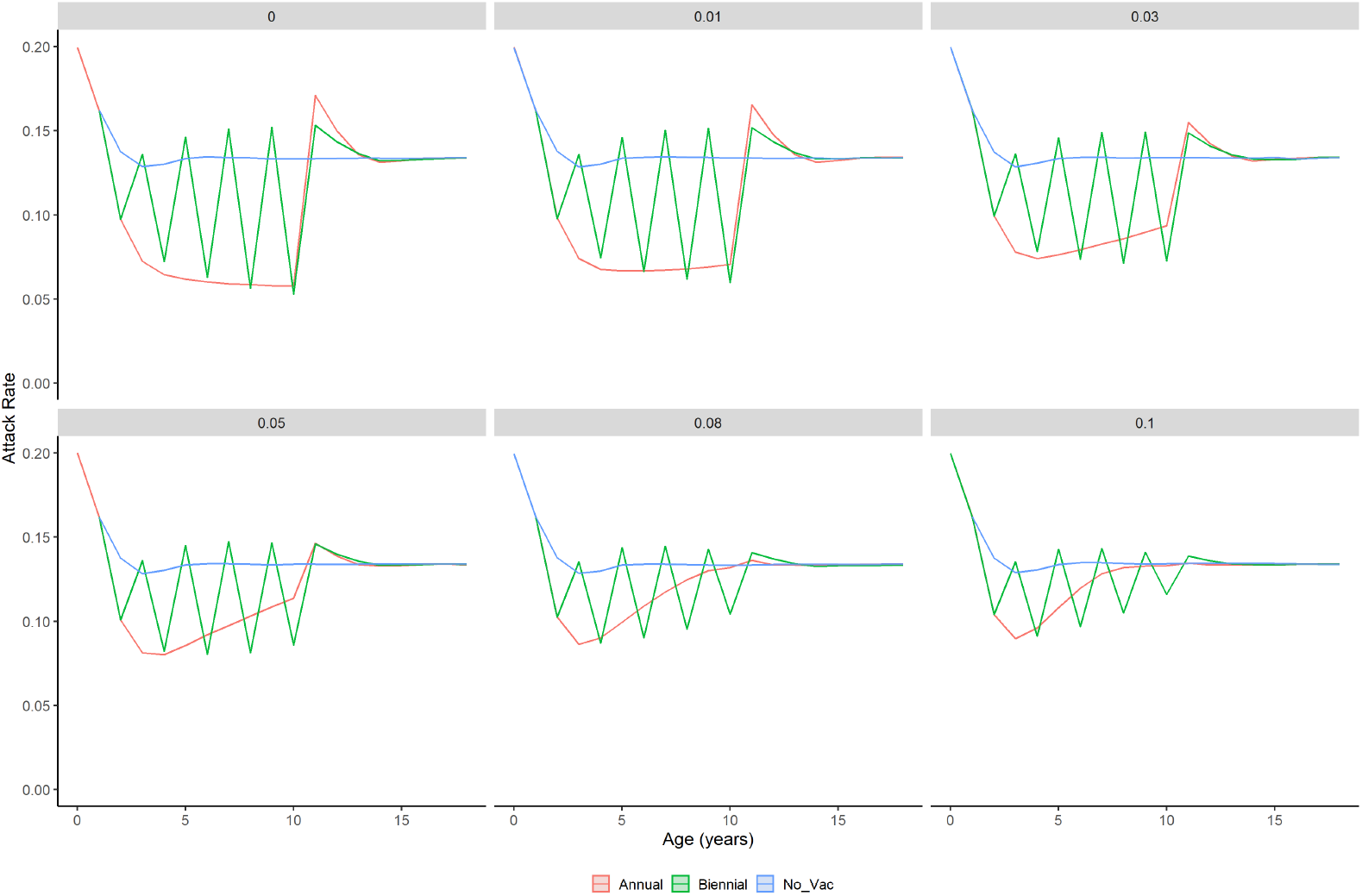
Average annual attack rates in birth cohorts under different vaccination strategies: annual (red), biennial (green), and no vaccination (blue) for different exposure penalties. Lines/shading indicate mean 95% percentiles from 1000 simulations. Children aged 2 to 10 years were vaccinated.

**Table 2.**
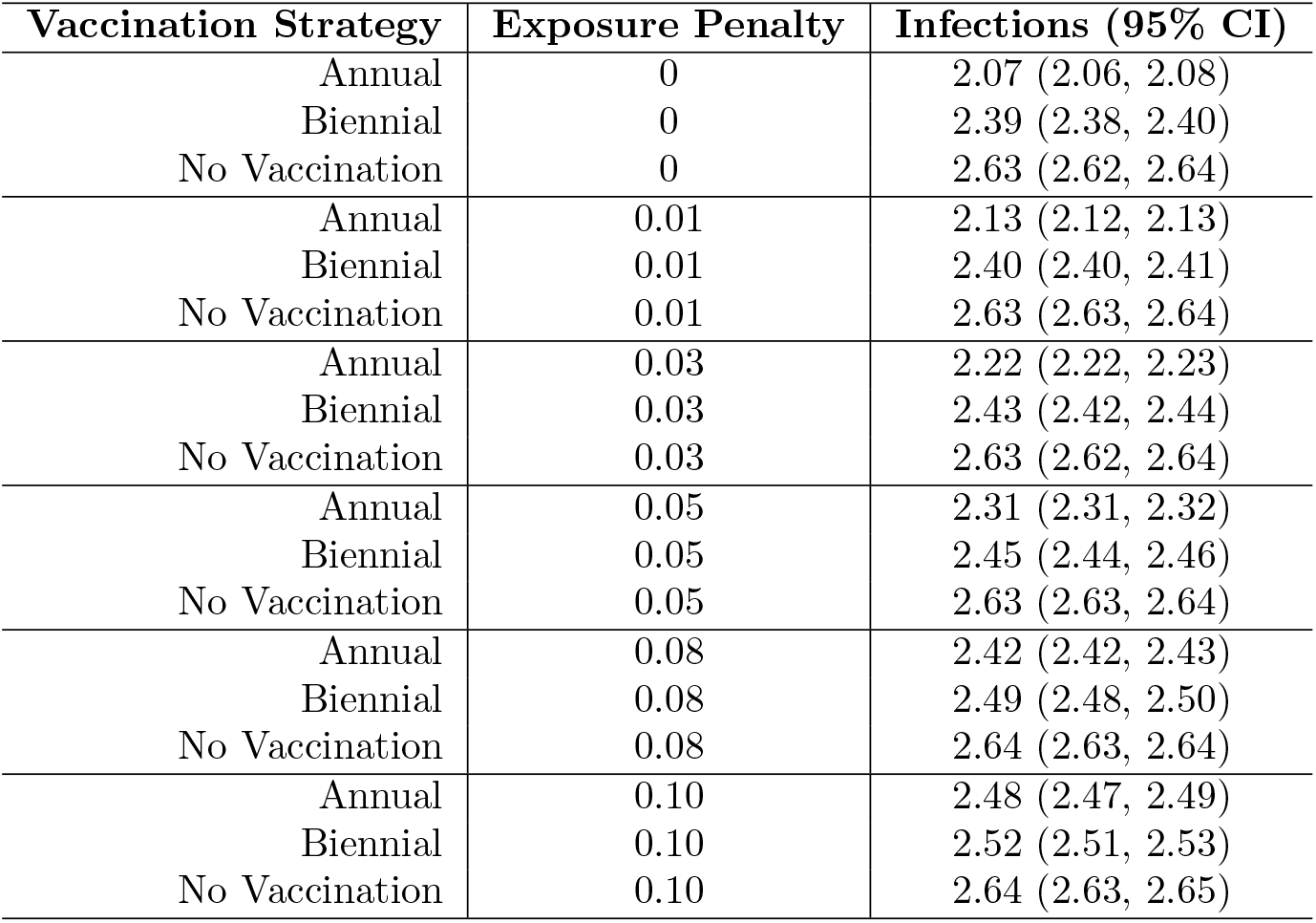
Number of expected childhood infections and 95% confidence intervals by vaccination strategy.

Next, we used the model to compare repeated annual vaccination and repeated biennial vaccination with respect to attack rates and expected number of infections up to the age of 18 (Fig 2, Table 1). At ages in which the vaccine was offered in both vaccination strategies, annual vaccination resulted in lower or similar attack rates compared to biennial vaccination under our baseline scenario (Fig 2A). For ages in which the vaccine was not offered under the biennial strategy, attack rates increased to similar or higher rates as that expected from the no vaccination strategy. Additionally, as we saw under the annual vaccination strategy, the attack rates under the biennial strategy increased after the oldest age of vaccination (age 11). Biennial vaccination resulted in 2.38 (2.37, 2.40) infections up to the age of 18 compared to 2.07 (2.06, 2.08) infections under annual vaccination and 2.64 (2.63, 2.66) under no vaccination (Table 1). When looking specifically at the years during the vaccination program (ages 2-10) we found that biennial vaccination resulted in an average of 0.889 (0.887, 0.892) infections compared to 0.568 (0.567, 0.570) infections under the annual vaccination program, and 1.20 (1.20, 1.20) infections under no vaccination.

As we observed with annual vaccination, the benefit of repeated biennial vaccination was sensitive to our assumption about the immunological exposure penalty. We found that as exposure penalty increased, the benefit of biennial vaccination decreased (Fig 3). Attack rates increased in later years of childhood under biennial vaccination (in ages in which the vaccine was offered); however, they increased more slowly than under annual vaccination, resulting in lower attack rates under biennial vaccination compared to annual vaccination later in childhood. The expected number of childhood infections up to age 18 under biennial vaccination ranged from 2.39 (2.38, 2.40) with no exposure penalty to 2.52 (2.51, 2.53) with the maximum exposure penalty (Table 2). For every value of exposure penalty, annual vaccination resulted in fewer childhood infections compared to biennial vaccination; however, the difference between the number of childhood infections from the two strategies decreased as exposure penalty increased. For example, with no exposure penalty, annual vaccination resulted in 2.07 (2.06, 2.08) infections compared to 2.38 (2.37, 2.40) for a difference of 0.31 infections, whereas with an exposure penalty of 0.1 the difference decreased to 0.04 infections (annual: 2.48 (2.47, 2.49), biennial: 2.52 (2.51, 2.53). Regardless of exposure penalty, both annual and biennial vaccination strategies resulted in fewer childhood infections compared to the no vaccination strategy.

When we relaxed our baseline assumptions and calculated the difference in expected numbers of childhood infections under annual vaccination versus no vaccination for various combinations of model parameter values, we found that annual vaccination resulted in fewer or similar childhood infections compared to no vaccination regardless of model parameter values (Fig 4A). The differences in expected infections with 95% confidence intervals that did not include zero ranged from −0.78 (−0.79, −0.77) to −0.01 (−0.02, 0.00), where difference is defined as the number of childhood infections under annual vaccination minus number of childhood infections under no vaccination. To assess how each model parameter impacted the difference in childhood infections, we created bivariate scatter plots for each combination of model parameters (Fig. S0). We found that vaccine effectiveness and exposure penalty had the greatest impact on vaccine strategy performance. Unsurprisingly, when vaccine effectiveness was low, the difference in expected childhood infections between the two vaccine strategies was small.

**Fig 4.**
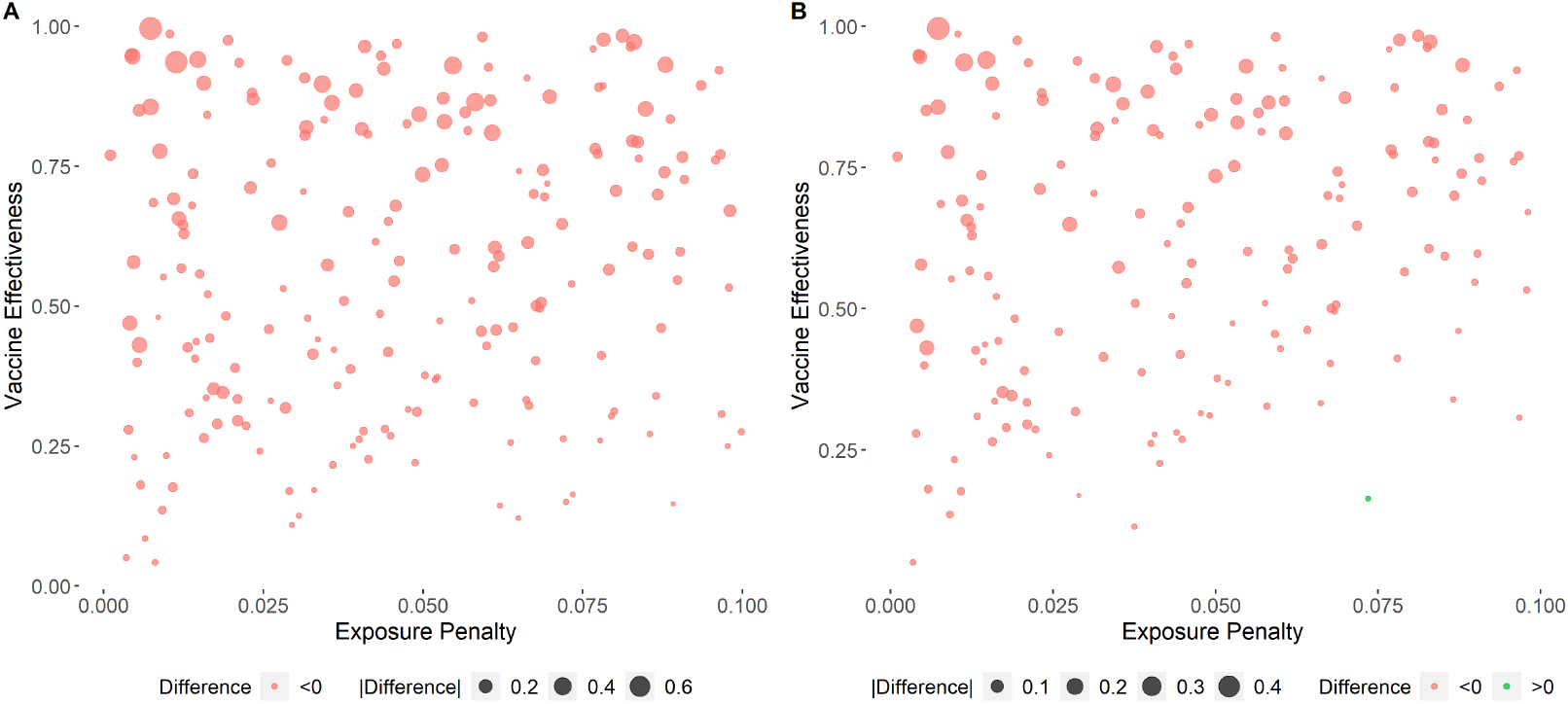
Bivariate scatter plot of vaccine effectiveness by exposure penalty for A) annual vs. no vaccination and B) annual vs. biennial. Points indicate difference in childhood infections between annual vaccination and either no vaccination (A) or biennial vaccination (B). Red indicates differences *<* 0 (indicating that annual vaccination resulted in fewer childhood infections). Green indicates differences *>* 0 (indicating that biennial vaccination resulted in fewer childhood infections than annual vaccination). The size of the dot indicates the magnitude of difference.

The differences in number of childhood infections under biennial vaccination versus annual vaccination ranged from −0.46 (−0.47, −0.45) to 0.01 (0, 0.03) when we varied our model parameter values, where difference is defined as number of childhood infections under annual vaccination minus the number of childhood infections under biennial vaccination (Fig 4B). We found that the largest absolute differences occurred for high values of vaccine effectiveness, and that annual vaccination resulted in fewer infections than biennial vaccination. However, when vaccine effectiveness was low, there were only small differences in the number of childhood infections resulting from the two vaccination strategies. To assess how each model parameter impacted the difference in childhood infections, we created bivariate scatter plots for each combination of model parameters (Fig. S1). We found that vaccine effectiveness and exposure penalty were the strongest determinants of vaccine strategy performance.

Current UK policy is to vaccinate children aged 2 to 10 years for influenza [5]; however that policy is expected to be extended to children aged 11 to 16 years [46]. Thus, we performed a sensitivity analysis where we evaluated annual and biennial vaccination strategies when vaccination occurred from ages 2 to 16 years first under the baseline scenario and then under the same parameter combinations from Latin hypercube sampling in the main analysis. When vaccination was extended until age 16 under the baseline scenario, fewer childhood infections were observed than when vaccination only included those aged 2-10 years. Specifically, annual vaccination resulted in 1.61 (1.61, 1.62) expected childhood infections compared to 2.20 (2.19, 2.21) childhood infections under biennial vaccination. When model parameters were varied, we found that annual vaccination mostly outperformed biennial vaccination (Fig. S2). There was no clear pattern of model parameters in which biennial vaccination outperformed annual vaccination; however, there were more parameter combinations for which biennial outperformed annual (green dots). These parameter combinations all featured an exposure penalty greater than 0 and many also corresponded to a high vaccine effectiveness (Fig. S2A).

## Discussion

Our work confirms the value of annual influenza vaccination in children by considering the average number of infections per participant during their childhood. We found that an annual influenza vaccination program reduced childhood infections compared to no vaccination program. Differences in childhood infections were most pronounced during the years in which children received the vaccine (ages 2-10). We observed a 2-fold reduction in childhood infections from ages 2-10 under an annual vaccination program in children who were vaccinated at least half the time compared to no vaccination. Due to our baseline assumptions that the vaccine-induced immunity waned completely after one year and vaccine coverage was 44% [43], averaging over all ages diluted differences in expected numbers of infections. Regardless, the reduction of infections under an annual vaccination program were robust to different model assumptions and support current childhood influenza vaccination programs in several countries [5, 32, 47, 48].

We also considered a biennial vaccination program in which children are vaccinated every other year. We found that while annual vaccination averts more childhood infections than biennial vaccination, in most scenarios considered in this work, this difference is not large. Under a key assumption that repeated exposures reduce the immune response to subsequent exposures, similar benefits of vaccination may be obtained by implementing a biennial vaccination program. This is of particular importance to countries where annual vaccination programs are logistically- or cost-prohibitive and supports an alternative influenza vaccination program in children. However, we do not perform a cost-benefit analysis on the different vaccination programs as was done in de Boer *et al*. [49]. Our assertion that biennial vaccination may be more feasible for countries where an annual influenza vaccination program in children is cost-prohibitive is based on the simple fact that vaccination every other year requires half the amount of vaccines as an annual vaccination program. Therefore, before any country adopts a biennial vaccination program, a full cost-benefit analysis should be conducted.

Throughout our different scenarios we observed that annual risk of infection under vaccination was higher in the period immediately following the cessation of repeated vaccination. This is likely due to an increase of fully susceptible individuals following the end of the vaccination program. We assume in the baseline scenario that vaccine protection wanes completely after one year resulting in higher susceptibility following the end of the vaccination program. Once the newly unvaccinated individuals become infected and acquire immunity from infection, we observe the attack rate level out to no vaccination program levels. A similar phenomenon was seen in Backer *et al*. after the end of the vaccination program [8]. The increase in attack rate in off years within a biennial vaccination program is also likely due to an increase in susceptible individuals with little natural immunity because we assume the vaccine wanes completely after one year in the baseline scenario. Therefore, in off years, individuals participating in the vaccination program are just as likely to get infected as those who are not participating in the vaccination program. When the assumed rate of waning of vaccine-induced immunity is lower, there will be carry-over protection in between vaccination years within a biennial program, reducing the risk of infection in off years.

While this study provides a framework by which to assess different frequencies of vaccination, it is a theoretical modelling study. Due to the unavailability of longitudinal data of repeated exposures in birth cohorts, this model has not been fit to data; however, the parameter values used in this study are informed by the literature. Longitudinal data of birth cohorts which track infection and vaccination history are not routinely collected. In 2019 the National Institutes of Health funded two birth cohort studies to evaluate the immune response to children’s first exposures to influenza [50]. Once data from these and similar studies is made available, this model can be adapted to fit to data.

Tracking longitudinal time series of infection and vaccination over many years are essential to understand how the immune system responds to similar exposures over time and the duration of immunity. Here, we assume that the exposure penalty due to infection and vaccination are the same. The immune response to subsequent exposures is not well understood and exposure to natural infection and vaccination may impact the immune system’s response differently. Despite the current gaps in scientific understanding, it is plausible that such a penalty is a real phenomenon. Thus, it is important to consider whether current vaccination programs unnecessarily use up immunity. This study highlights the importance of determining the impact of repeated exposure on the immune response.

The approach taken here for children may be appropriate for studies of COVID-19 vaccination in all ages. As more data accumulate on the duration of protection from severe disease and the impact of evolutionary change, many countries may choose to opt for repeated vaccine boosters based on update target lineages. Should this occur, the same trade-offs will likely apply as have been demonstrated here. Crucially, longitudinal studies should be started as soon as possible so that the impact of repeated vaccination can be estimated.

We make several simplifying assumptions in this work. Specifically, we do not stratify our population by risk factor. Thus, we assume that the risk of infection is determined by prior exposure only and not an underlying condition. We don’t expect this simplification to impact our results. We also assume that vaccine effectiveness is the same for all individuals regardless of age. There is evidence that suggests that vaccines may be less effective in the elderly [51]. However, the focus of this modelling study is on children and thus, the assumed vaccine effectiveness in other age groups will not impact the results presented here. Additionally, vaccine characteristics such as waning immunity are unlikely to vary among the similarly aged individuals who are the focus group of this work. Therefore, we assume a fixed waning rate. Finally, we consider a model with only one circulating virus strain, thus we do not incorporate cross-reactivity of similar strains into our susceptibility model. A similar approach has been taken in other modelling studies [8, 16, 49].

In conclusion, we developed a model to assess the potential benefits of long-term annual repeated childhood vaccination. Our results demonstrate the benefit of annual childhood vaccination in preventing infections; however, in the presence of an exposure penalty that reduces the immune response to subsequent exposures, we found that a biennial vaccination program resulted in similar benefits. These results are the theoretical differences in direct effects of different vaccination strategies, and we have not attempted to quantify indirect effects, which formed a large component of prior modelling work to justify the annual vaccination policy in the UK [7, 52]. The differences we show for some scenarios are small, therefore the differences in indirect benefits are unlikely to be overwhelming. However, more thorough modelling work is required before the concepts presented here can be applied directly to policy.

## Data Availability

The open source package morevac is available on GitHub

https://github.com/kylieainslie/morevac

## Supporting information

**Fig. S0.**
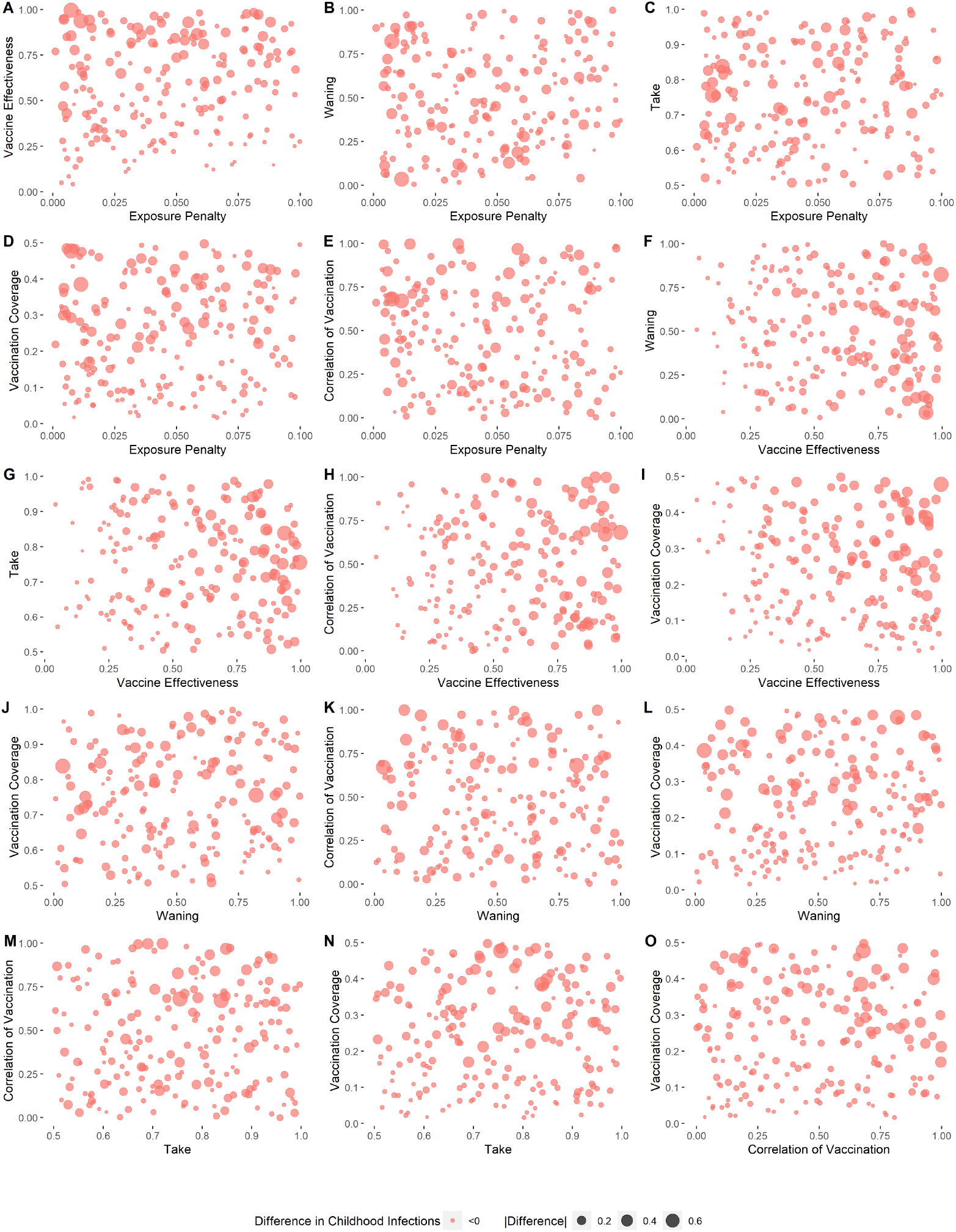
Bivariate scatter plots of average difference in childhood infections (annual - no vaccination) for every pair of model parameters when vaccination occurs from ages 2-10. Red indicates differences *<* 0 (annual results in fewer childhood infections). The size of the dot indicates the magnitude of difference.

**Fig. S1.**
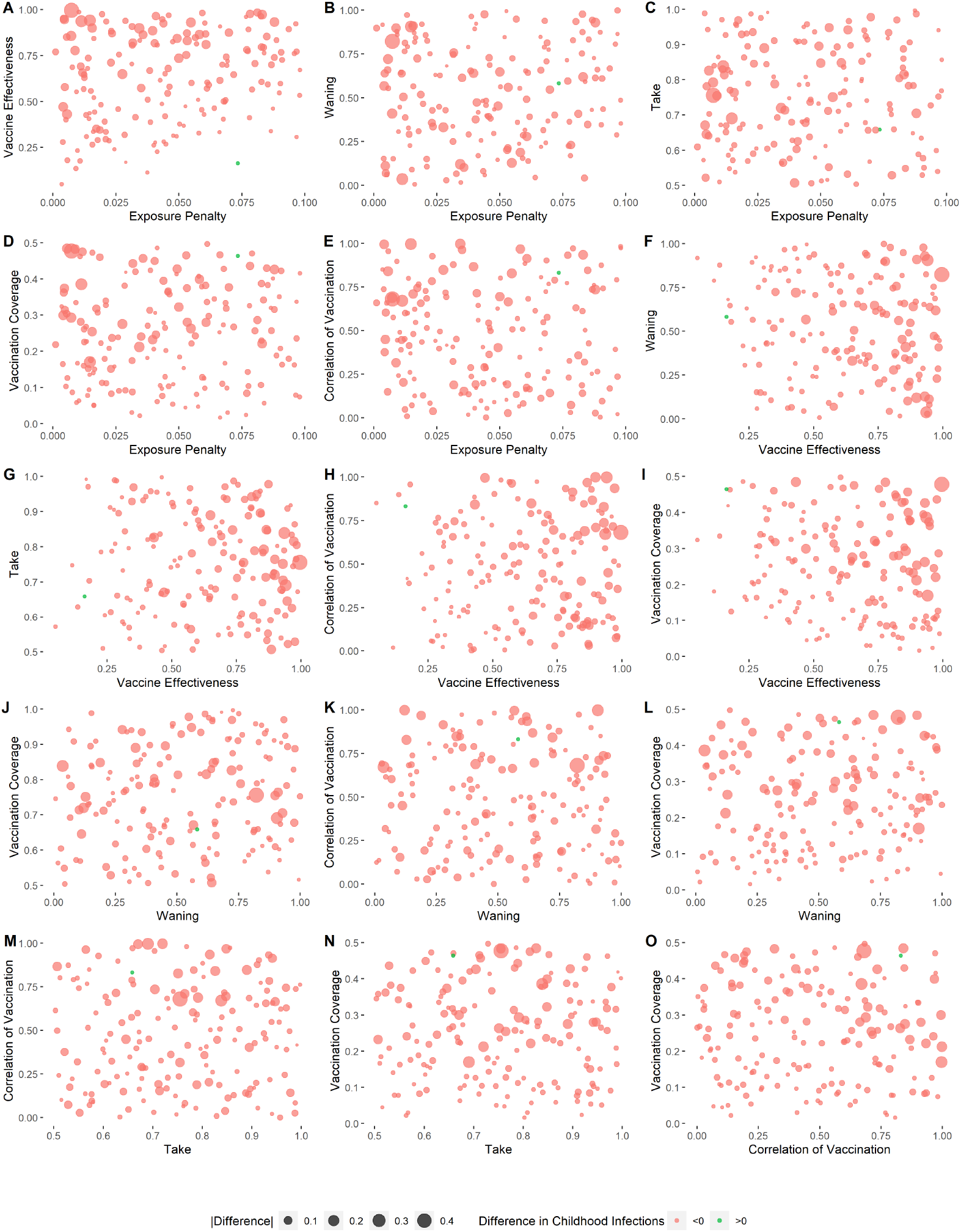
Bivariate scatter plots of average difference in childhood infections (annual - biennial) for every pair of model parameters when vaccination occurs from ages 2-10. Red indicates differences *<* 0, green indicates differences *>* 0. The size of the dot indicates the magnitude of difference. When the difference *<* 0 (red dots), annual vaccination results in less childhood infections compared to biennial vaccination. When the difference *>* 0 (green dots), biennial vaccination results in less childhood infections compared to annual vaccination.

**Fig. S2.**
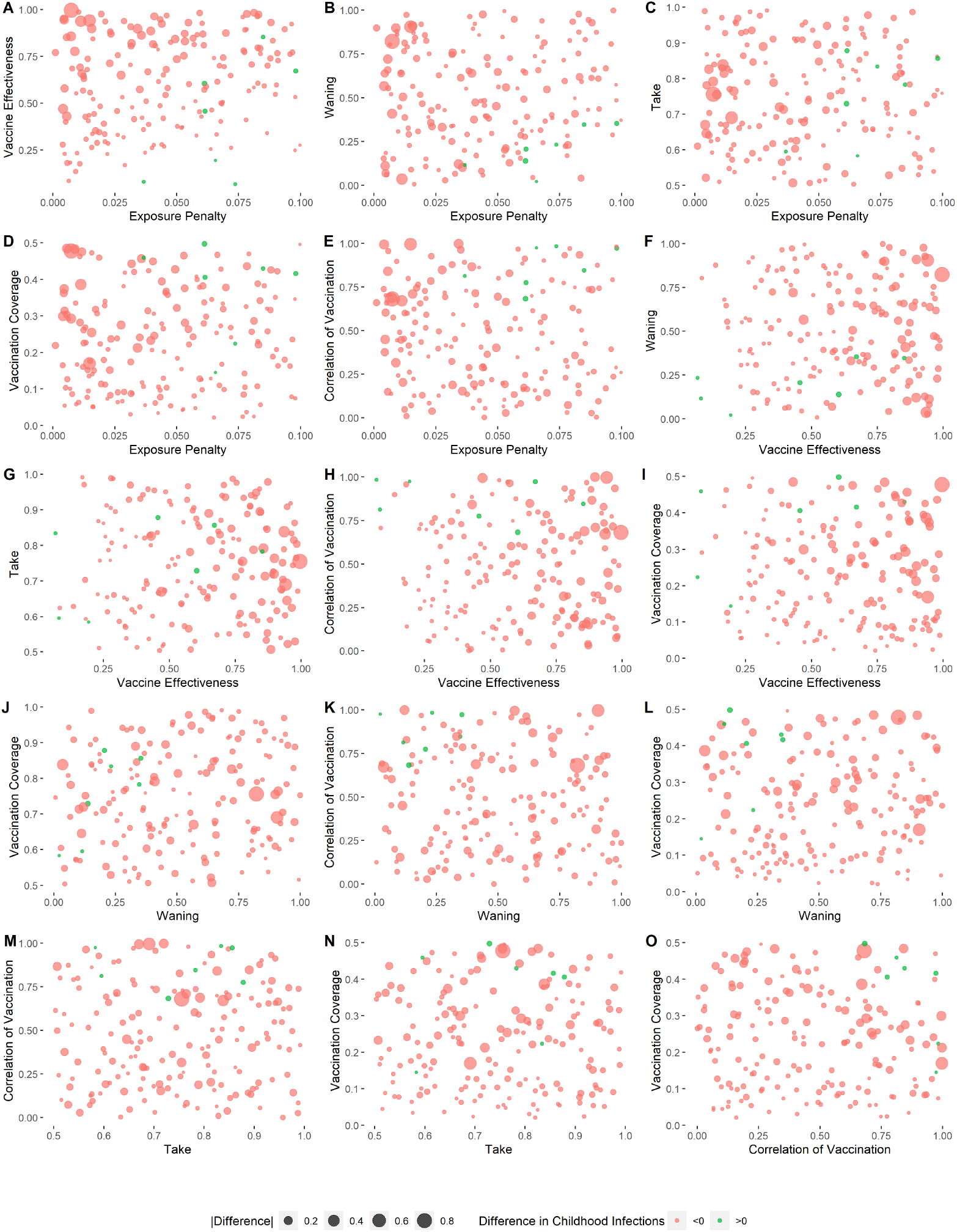
Bivariate scatter plots of average difference in childhood infections (annual - biennial) for every pair of model parameters when vaccination occurs from ages 2-16. Red indicates differences *<* 0, green indicates differences *>* 0. The size of the dot indicates the magnitude of difference. When the difference *<* 0 (red dots), annual vaccination results in less childhood infections compared to biennial vaccination. When the difference *>* 0 (green dots), biennial vaccination results in less childhood infections compared to annual vaccination.

**Fig. S3.**
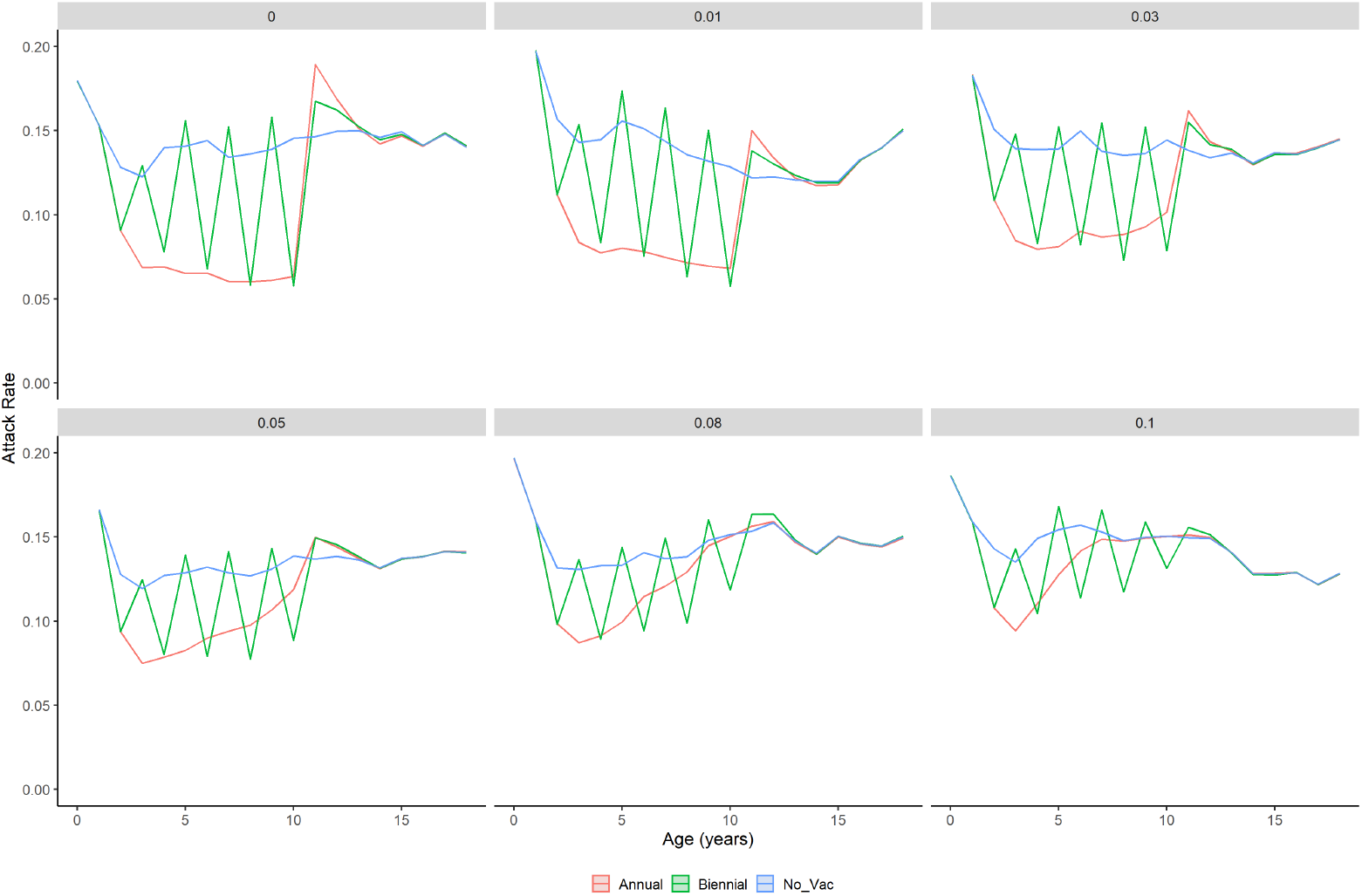
Average annual attack rates with time-varying force of infection in birth cohorts under different vaccination strategies: annual (red), biennial (green), and no vaccination (blue) for different values of exposure penalty. Lines/shading indicate mean 95% percentiles from 1000 simulations. Children aged 2 to 10 years were vaccinated.

**Table S1.** Number of expected childhood infections and 95% confidence intervals by vaccination strategy when force of infection varies by year.

| Vaccination Strategy | Exposure Penalty | Infections (95% CI) |
| --- | --- | --- |
| Annual | 0 | 2.07 (2.07, 2.08) |
| Biennial | 0 | 2.39 (2.39, 2.39) |
| No Vaccination | 0 | 2.63 (2.63, 2.64) |
| Annual | 0.01 | 2.12 (2.12, 2.13) |
| Biennial | 0.01 | 2.41 (2.40, 2.41) |
| No Vaccination | 0.01 | 2.63 (2.62, 2.63) |
| Annual | 0.03 | 2.22 (2.22, 2.22) |
| Biennial | 0.03 | 2.43 (2.42, 2.43) |
| No Vaccination | 0.03 | 2.63 (2.62, 2.63) |
| Annual | 0.05 | 2.31 (2.31, 2.32) |
| Biennial | 0.05 | 2.46 (2.45, 2.46) |
| No Vaccination | 0.05 | 2.63 (2.63, 2.64) |
| Annual | 0.08 | 2.43 (2.43, 2.44) |
| Biennial | 0.08 | 2.49 (2.49, 2.50) |
| No Vaccination | 0.08 | 2.63 (2.62, 2.63) |
| Annual | 0.10 | 2.48 (2.47, 2.48) |
| Biennial | 0.10 | 2.52 (2.51, 2.52) |
| No Vaccination | 0.10 | 2.63 (2.63, 2.64) |

